# Pathological uptake with 18-Fluorocholine versus 99mTc-MIBI in the location of the parathyroid glands in hyperparathyroidism. Systematic review and meta-analysis

**DOI:** 10.1101/2020.07.25.20161927

**Authors:** José Luis Pardal Refoyo, Pilar Tamayo Alonso, Sofía Ferreira Cendón, Esther Martín Gómez

## Abstract

**Introduction:** The location of the pathological parathyroid glands in hyperparathyroidism is usually carried out by means of 99mTc-sestamibi scintigraphy, which increases its precision by adding the ultrasound examination. The non-localization of the parathyroid glands increases the difficulties for surgical removal. To increase the detection of pathological glands, other radioactive tracers are used, such as methionine, fluorocholine or 18F-flurpiridaz.

**Objective:** To establish if PET / CT with 18-Fluorocholine in patients with hyperparathyroidism increases the number of uptakes compared to the 99mTc-sestamibi scan.

**Method:** Systematic review and meta-analysis. Two subgroups were analyzed. Subgroup 1: trials comparing both techniques as an initial exploration. Thirteen studies including 1131 examinations were selected (596 PET / CT with 18-Fluorocholine vs. 535 scintigraphy with 99mTc-sestamibi). Meta-analysis was performed following the random effects model and the odds ratio was calculated. Subgroup 2: studies that include 18-Fluorocholine as a rescue examination in patients with a previous negative study with a 99mTc-sestamibi scan. 17 articles including 412 examinations with 359 patients in which there was at least one uptake were selected. Meta-analysis of the prevalence of the number of patients in whom there was at least one uptake was performed using the random effects model.

**Results:** Subgroup 1: The number of patients in which at least one uptake occurs is significantly higher with the 18-Fluorocholine examinations (OR 4.264, 95% CI 2.400-7.577). The prevalence of uptake with 18-Fluorocholine is 0.91 [0.86, 0.95] and with sestamibi 0.68 [0.56, 0.80]. Subgroup 2: the prevalence of uptake among patients with previous negative MIBI studies was 0.90 [0.87, 0.94]. The probability of detection of both techniques in this group reaches 0.98. Publication bias in the meta-analyzes is low.

**Discussion:** 18-Fluorocholine protocols provide higher precision, clearer images, with faster acquisition as well as being readily available for most PET / CT centers.

**Conclusion:** The PET / CT study with 18-Fluorocholine can be recommended as a study for the location of pathological parathyroid glands after studies with negative MIBI.

## Introduction

The location of the pathological parathyroid glands in hyperparathyroidism is usually performed by SPECT/TC scan with 99mTc-sestamibi (MIBI) which increases their accuracy when associating ultrasound scanning. The non-location of the parathyroid glands increases difficulties for surgical removal. Other radioactive tracers such as methionine or fluorocholine are used to increase the detection of pathological glands [1–8].

The usefulness of PET/TC with 18-Fluorocholine (CH) was established after incidental findings of pathological parathyroid in males tracking for prostate cancer. In 2012 Mapelli et al., in 2013 Quak et al. and, in 2014 Hodolic et al. described cases of incidental uptake in parathyroids in explorations with F-18-fluorocholine in males with prostate cancer and proposed it as a study in case of negative MIBI [9–11]. Subsequent studies have confirmed this indication [12–14].

The accuracy of nuclear scans in the location of pathological parathyroid glands in hyperparathyroidism has been widely published and debated in the literature and the absence of localization poses a greater difficulty in surgical treatment so research is continued to establish a diagnostic strategy that offers the highest number of true positives and fewer false negatives. The objective of this review is to know whether PET/TC with 18-Fluorocholine in patients with hyperparathyroidism increases the number of catches compared to the scan with 99mTc-MIBI by bibliographic review.

## Method

The following research PICO question was asked: In patients with primary hyperparathyroidism [patient] the location of the pathological glands [intervention] by PET/TC with 18-Fluorocholine (CH) versus SPECT/TC with 99mTc-MIBI (MIBI) [comparison] does it offer more detections with suspected pathology? [outcome].

Systematic review in the PubMed (https://pubmed.ncbi.nlm.nih.gov/), WoS (http://wos.fecyt.es/), ClinicalTrials (https://clinicaltrials.gov/) and Cochrahe (https://www.cochranelibrary.com/) databases with closure on December 17, 2019 following the guidelines of the PRISMA group [15] and Cochrane method [16] with the search descriptors and strategies summarized in Figure 1.

**Figure 1.**
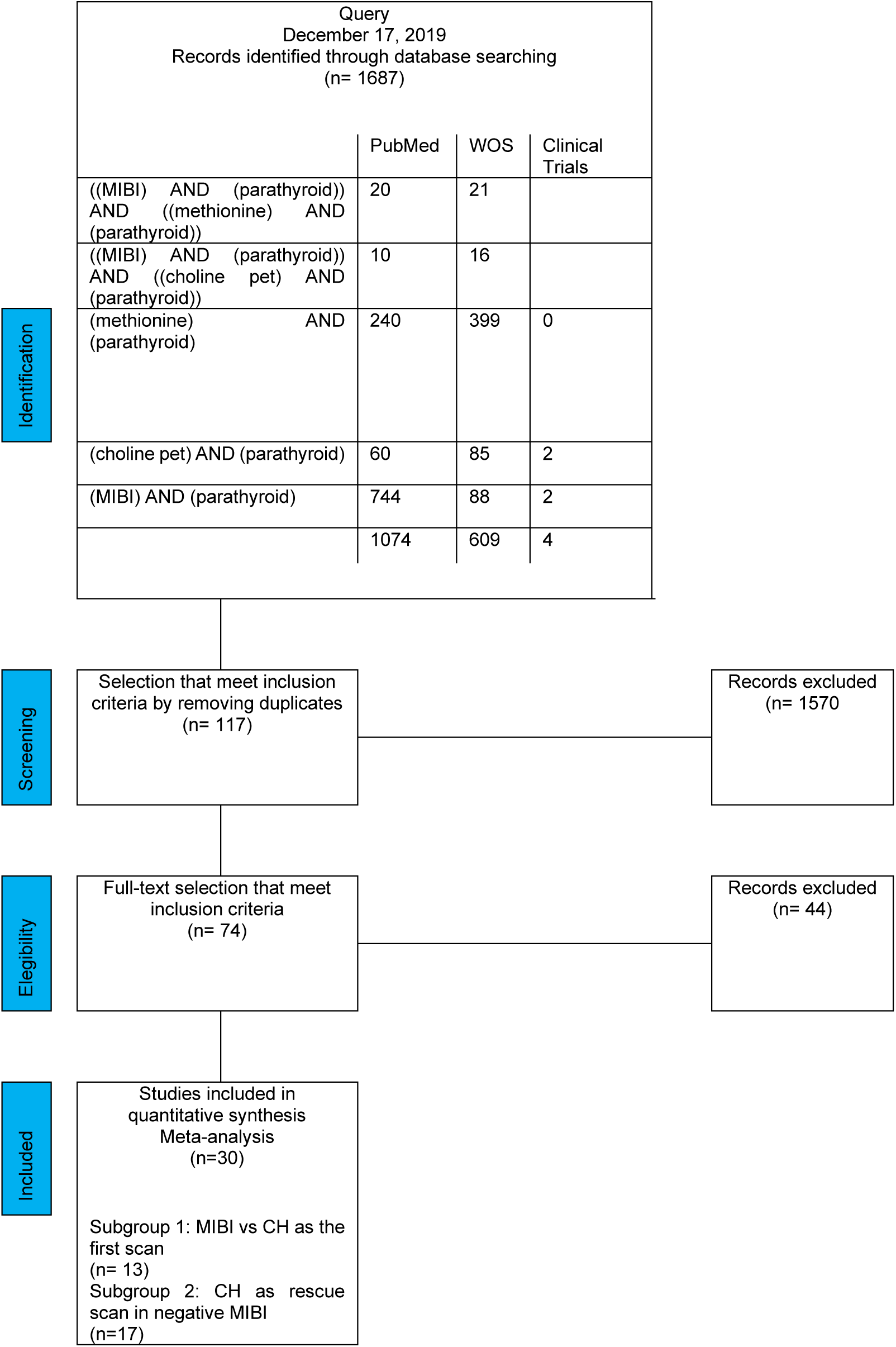
Bibliographical search and literature selection strategies.

Inclusion criteria: studies including patients with hyperparathyroidism undergoing scan with 99mTc-Sesta-MIBI (MIBI) and 18F-choline PET/CT (CH) as first scan (subgroup 1) and patients with study 99mTc-Sesta-MIBI (MIBI) negative in the first scan to be performed second rescue scan with 18F-choline PET/CT (CH) in which there was at least a significant scan uptake (subgroup 2). Articles indicating the number of patients scanned and the number of patients in whom there was at least a positive uptake in the neck or mediastinum and those in which there was no uptake or this nor was conclusive evaluated with SUVmax measurement (maximum standardized absorption value).

Exclusion criteria: clinical cases, conferences, conferences with published data or parathyroid cancer series exclusively.

Each article obtained the total number of patients with scans performed and calculated the prevalence of patients in which there was at least one uptake (positive uptake) or there was no uptake, or this was doubtful (negative uptake).

Two subgroups were analyzed:

Subgroup 1: essays comparing both techniques (CH *vs*. MIBI) as an initial scan.

Meta-analysis of dicotomical models was performed following the random effects model, the *odds ratio* and the publication bias were calculated.

Subgroup 2: studies that include CH as a rescue scan in patients with prior negative study with MIBI scan.

Meta-analysis of the prevalence of the number of patients in which there was at least one uptake was performed using the random effects model and the publication bias was evaluated.

The statistical program JAMOVI 1.2.22 (The jamovi project 2020, https://www.jamovi.org) was used.

Study limitations

- Only patients where at least one scan uptake was detected.
- The number of uptakes per patient was not evaluated.
- Patients with primary hyperparathyroidism are included but in some cases the diagnosis is not clear.
- Correlation of catchments with surgical findings was not sought to assess the accuracy of the tests.
- It is assumed that the scan uptake increases the probability of surgical location.

## Results

Subgroup 1:

13 studies including 1131 scans (596 PET/TC with 18-Fluorocholine vs. 535 SPECT/TC scan with 99mTc-sestamibi) [14,17–28] were selected. See Table 1.

**Table 1.** Subgroup 1: MIBI vs CH as the first scan. Results table.

|  | CH PET/TC |  |  | 99mTc-sestamibi SPECT/CT |  |  |
| --- | --- | --- | --- | --- | --- | --- |
| study name | N | CAPTURES | NO CAPTURE | N | CAPTURES | NO CAPTURE |
| Thansser 2017 [17] | 54 | 52 | 2 | 54 | 42 | 12 |
| Orevi 2014 [21] | 40 | 37 | 3 | 40 | 33 | 7 |
| Beheshti 2018 [22] | 82 | 76 | 6 | 82 | 50 | 32 |
| Araz 2018 [24] | 35 | 30 | 5 | 35 | 28 | 7 |
| Bossert 2019 [25] | 34 | 24 | 10 | 34 | 5 | 29 |
| Araz 2018 2 [23] | 43 | 42 | 1 | 43 | 35 | 8 |
| Imamovic 2016 [26] | 41 | 39 | 2 | 41 | 27 | 14 |
| Hocevar 2017 [27] | 151 | 126 | 25 | 90 | 74 | 16 |
| Kluijfhout 2016 [28] | 44 | 34 | 10 | 44 | 14 | 30 |
| Michaud 2015 [14] | 17 | 15 | 2 | 17 | 15 | 2 |
| Khafif 2019 [19] | 19 | 16 | 3 | 19 | 14 | 5 |
| Lezaic 2014 [20] | 24 | 24 | 0 | 24 | 20 | 4 |
| Michaud 2014 [18] | 12 | 11 | 1 | 12 | 7 | 5 |
|  | 596 | 526 | 70 | 535 | 364 | 171 |

The number of patients with at least one uptake is significantly higher with 18-Fluorocholine scans (OR 4,264, CI 95% 2,400-7,577). The prevalence of uptakes with 18-Fluorocholine is 0.91 [0.86, 0.95] and with MIBI of 0.68 [0.56, 0.80]. See Figures 2 and 3. The publication bias is moderate (I2-55.35%). See Figure 4.

**Figure 2.**
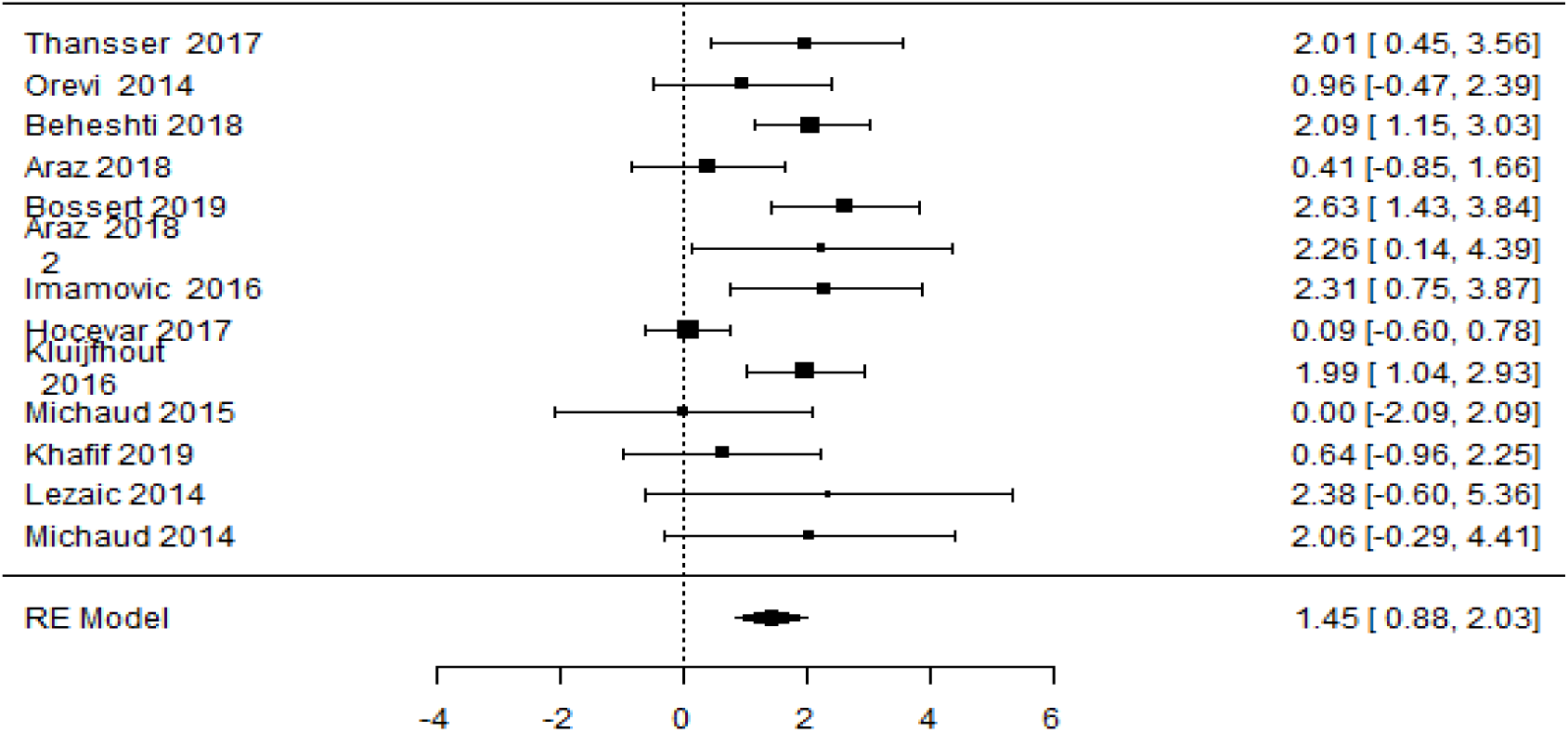
Subgroup 1: MIBI vs CH as the first scan. Forest plot.

**Figure 3.**
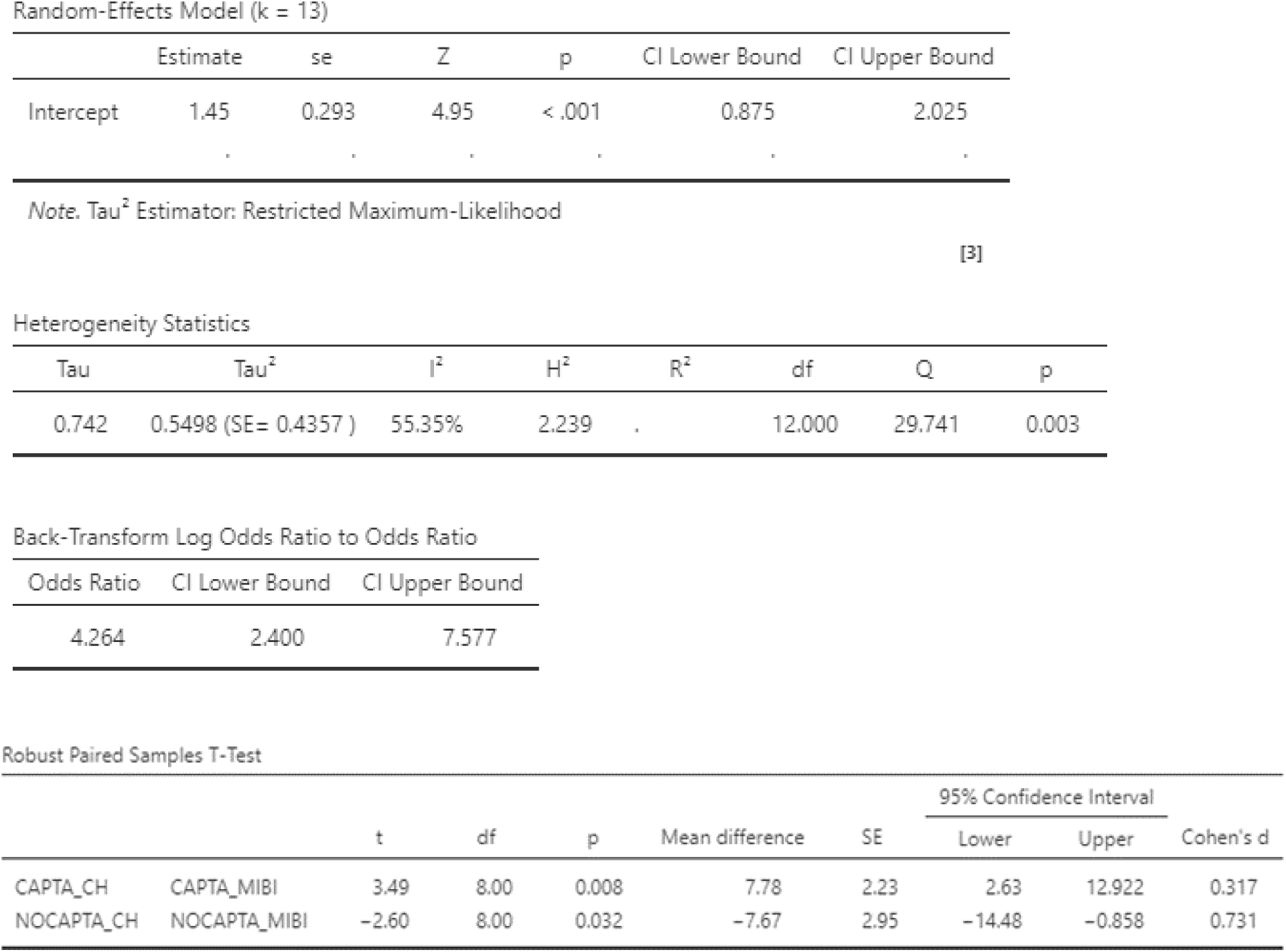
Statistical heterogeneity.

**Figure 4.**
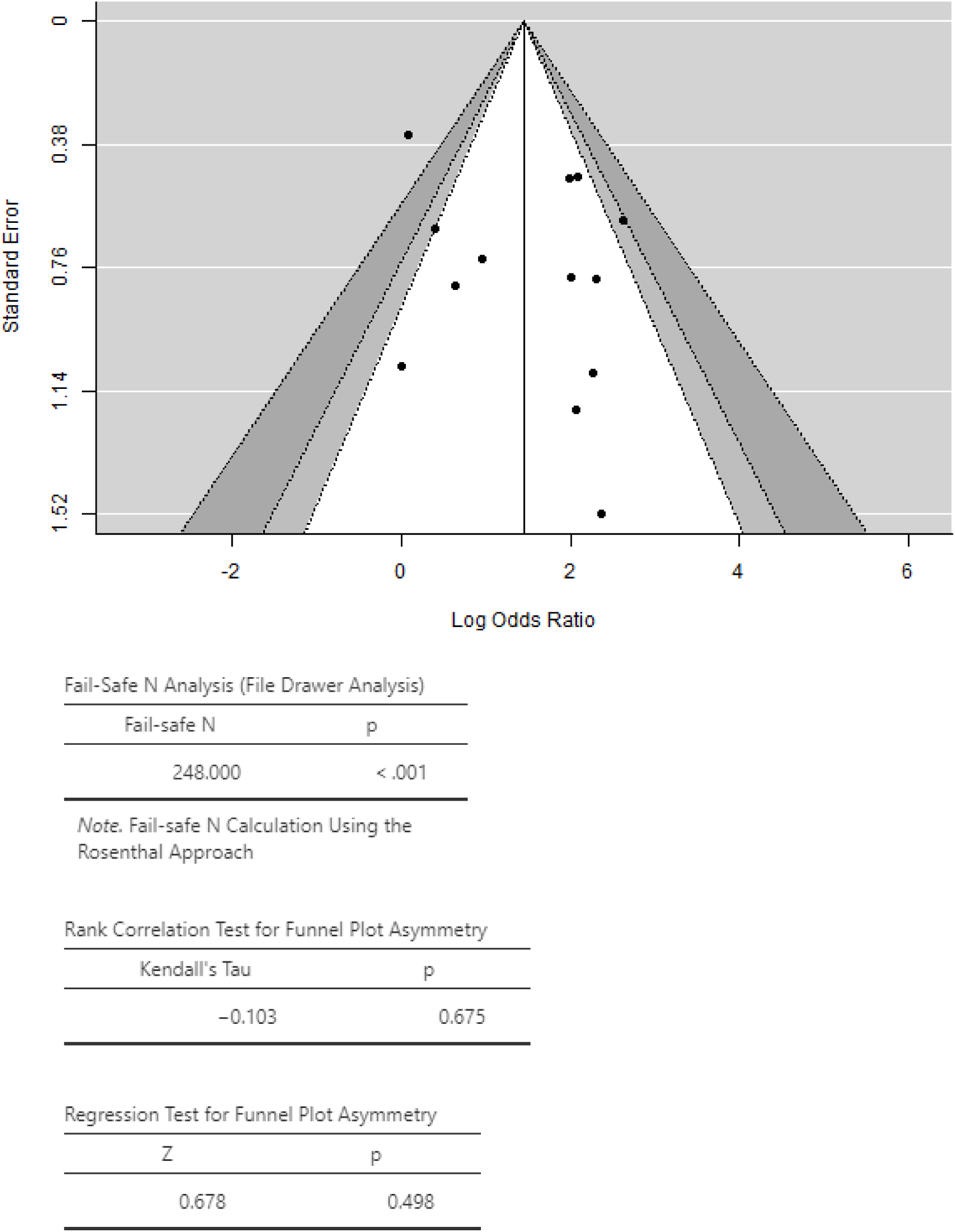
Subgroup 1: MIBI vs CH as the first scan. Funnel plot and publication bias

Subgroup 2:

17 articles including 412 scans were selected with at least 359 patients in which there was at least one uptake [17,21,23,29–41]. See Table 2.

**Table 2.** Subgroup 2: CH as a rescue scan in negative MIBI. Results table.

| Study name | N | CAPTURES | NO CAPTURE |
| --- | --- | --- | --- |
| Piccardo 2019 [35] | 44 | 32 | 12 |
| Thansser 2017 [17] | 11 | 11 | 0 |
| Orevi 2014 [21] | 7 | 7 | 0 |
| Zajickova 2018 [36] | 13 | 12 | 1 |
| Kluijfhout 2017 [37] | 10 | 9 | 1 |
| Quak 2018 [38] | 25 | 22 | 3 |
| Morales 2018 [39] | 49 | 42 | 7 |
| Bera 2018 [40] | 29 | 26 | 3 |
| Amadou 2019 [41] | 29 | 28 | 1 |
| Liu 2019 [29] | 87 | 70 | 17 |
| Christakis 2019 [30] | 12 | 12 | 0 |
| Collaud 2019 [31] | 7 | 6 | 1 |
| Grimaldi 2018 [32] | 27 | 24 | 3 |
| Huber 2018 [33] | 26 | 25 | 1 |
| Kluijfhout 2015 [37] | 5 | 4 | 1 |
| Treglia 2016 [34] | 23 | 21 | 2 |
| Araz 2018 2 [23] | 8 | 8 | 0 |
|  | 412 | 359 | 53 |

The prevalence of uptakes among patients with prior negative MIBI studies was 0.90 [0.87, 0.94]. The probability of detection of both techniques in this group reaches 0.98. See Figures 5 and 6. The publication bias in meta-analysis is low (I2-34.5%). See Figure 7.

**Figure 5.**
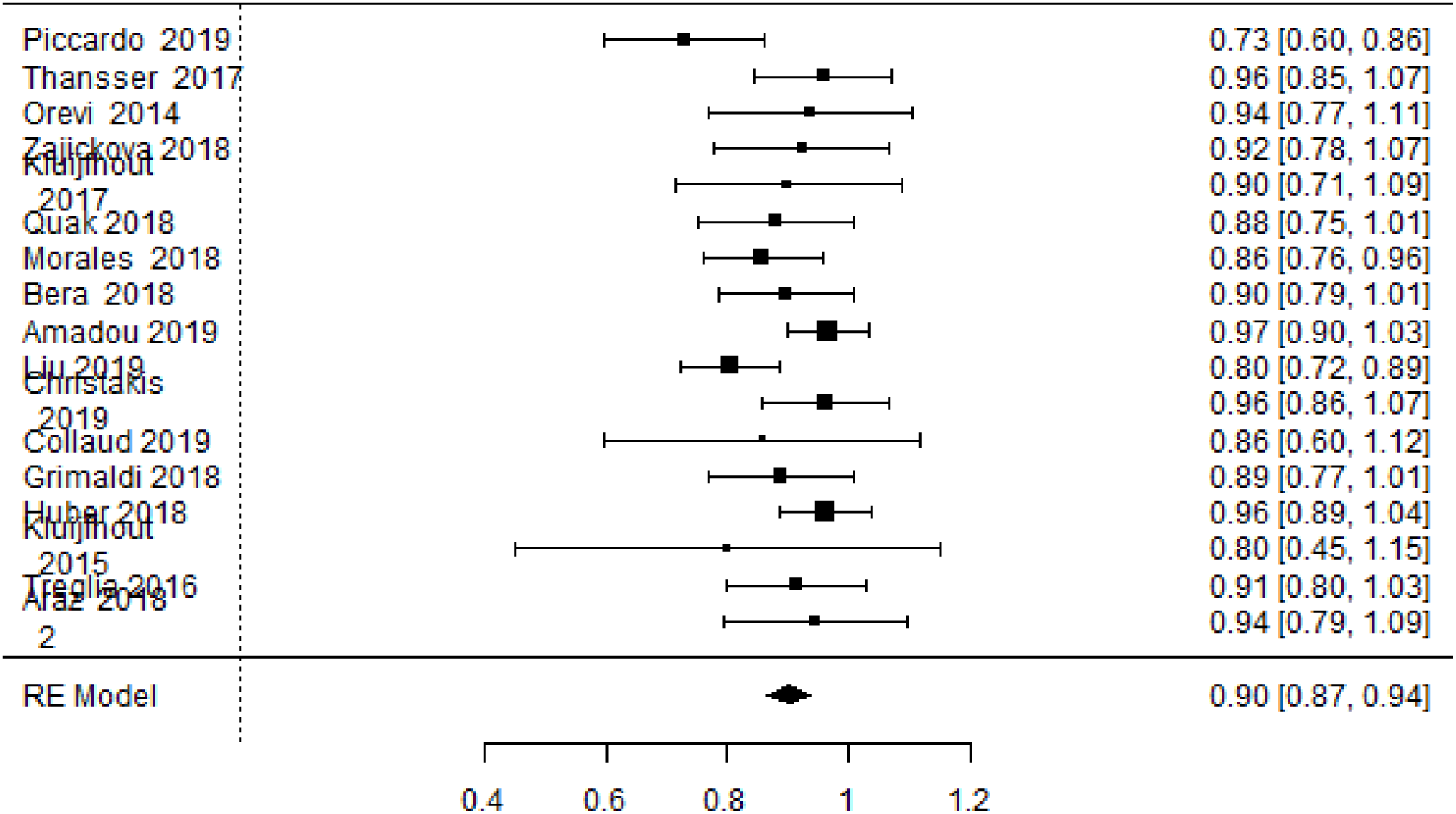
Subgroup 2: CH as a rescue scan in negative MIBI. Forest plot.

**Figure 6.**
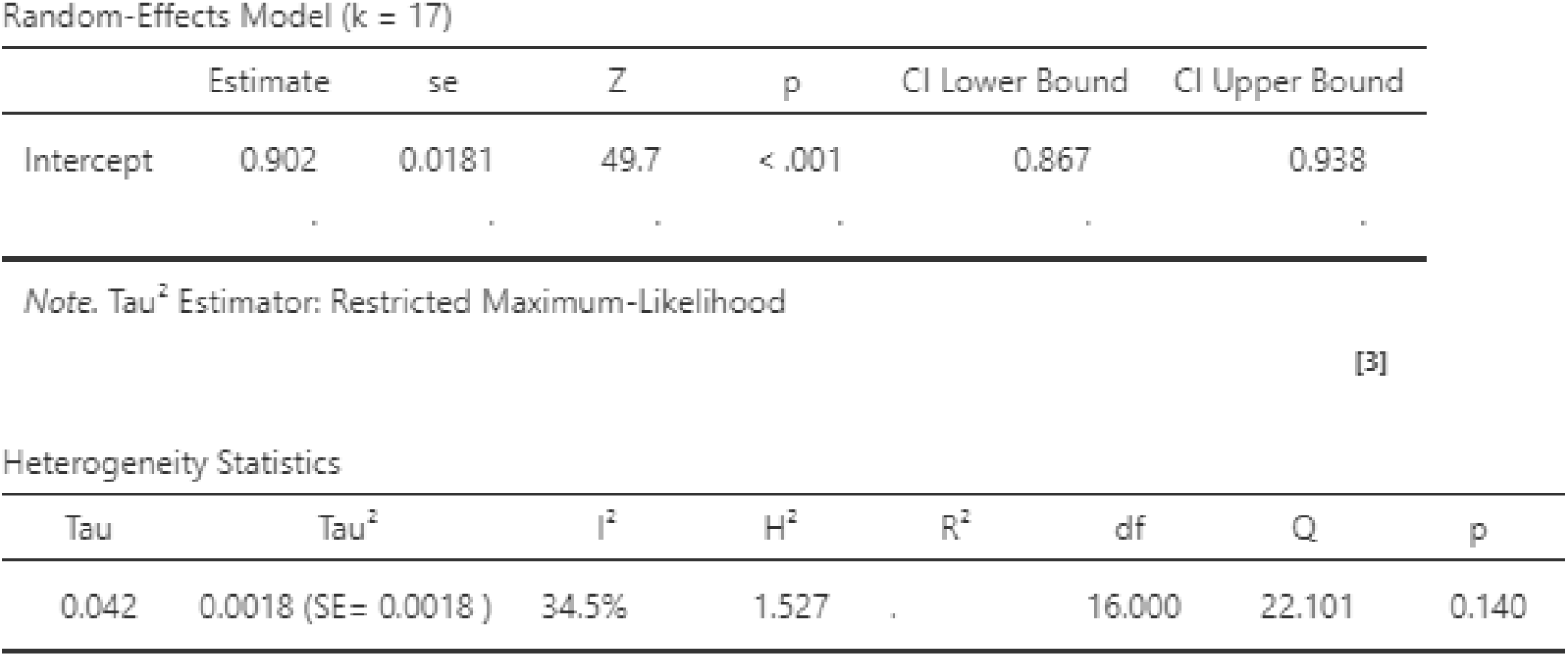
Statistical heterogeneity

**Figure 7.**
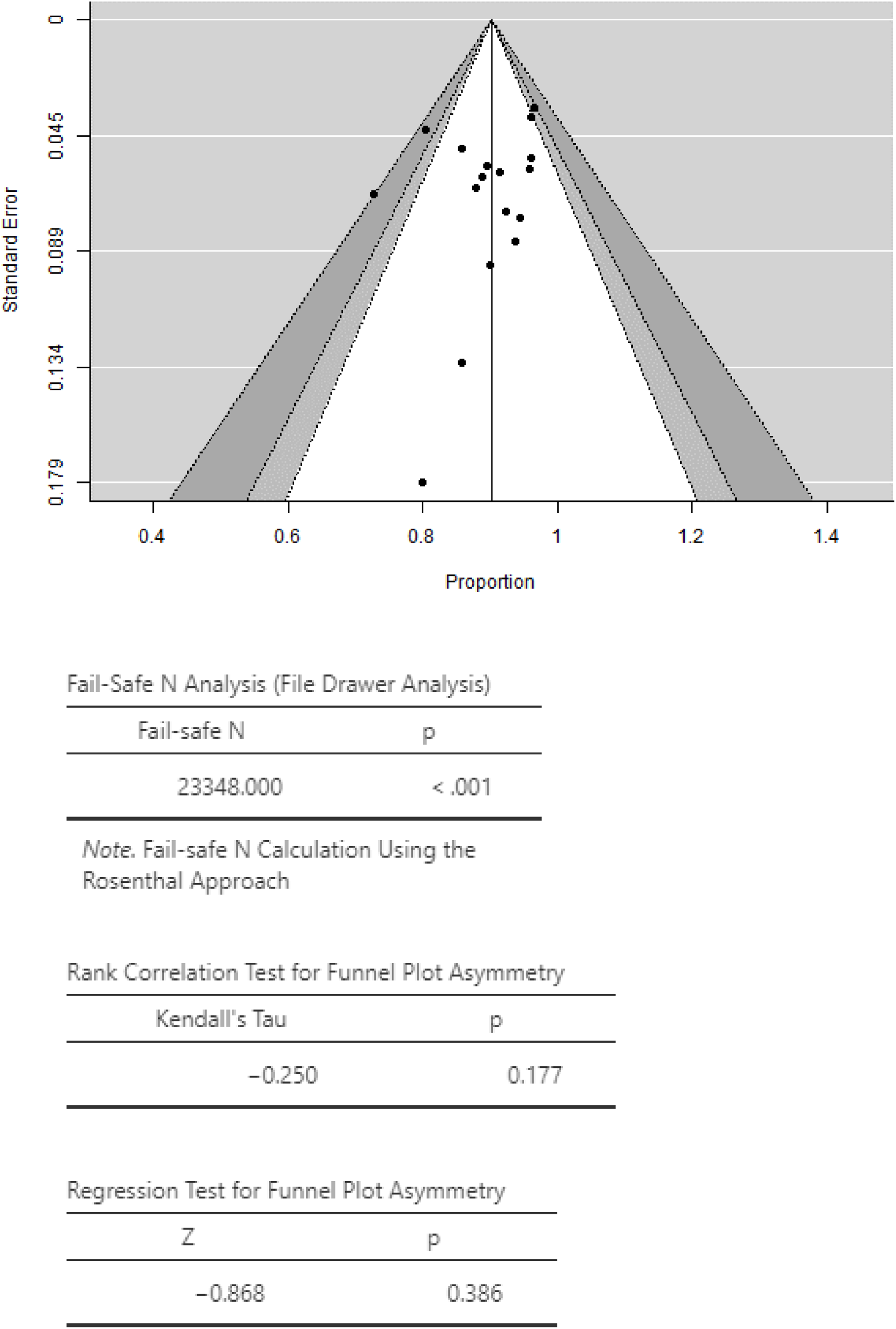
Subgroup 2: CH as a rescue scan in negative MIBI. Funnel plot and publishing bias.

## Discussion

Currently, the definitive treatment of hyperparathyroidism is the surgical removal of the gland or pathological glands. Negative localization studies make surgical treatment difficult.

In both the selected articles in our meta-analysis and in previous meta-analysis, CH localization studies have shown greater ability to locate pathological glands [34,42–45] than MIBI or methionine [46–48] in both first exploration and rescue scan [31,49].

The probability of uptake with CH is above 0.9 on both first scans and scans performed after negative MIBI study (in these cases the combination of both techniques reaches a prevalence of uptake of 0.97). See Figure 8.

**Figure 8.**
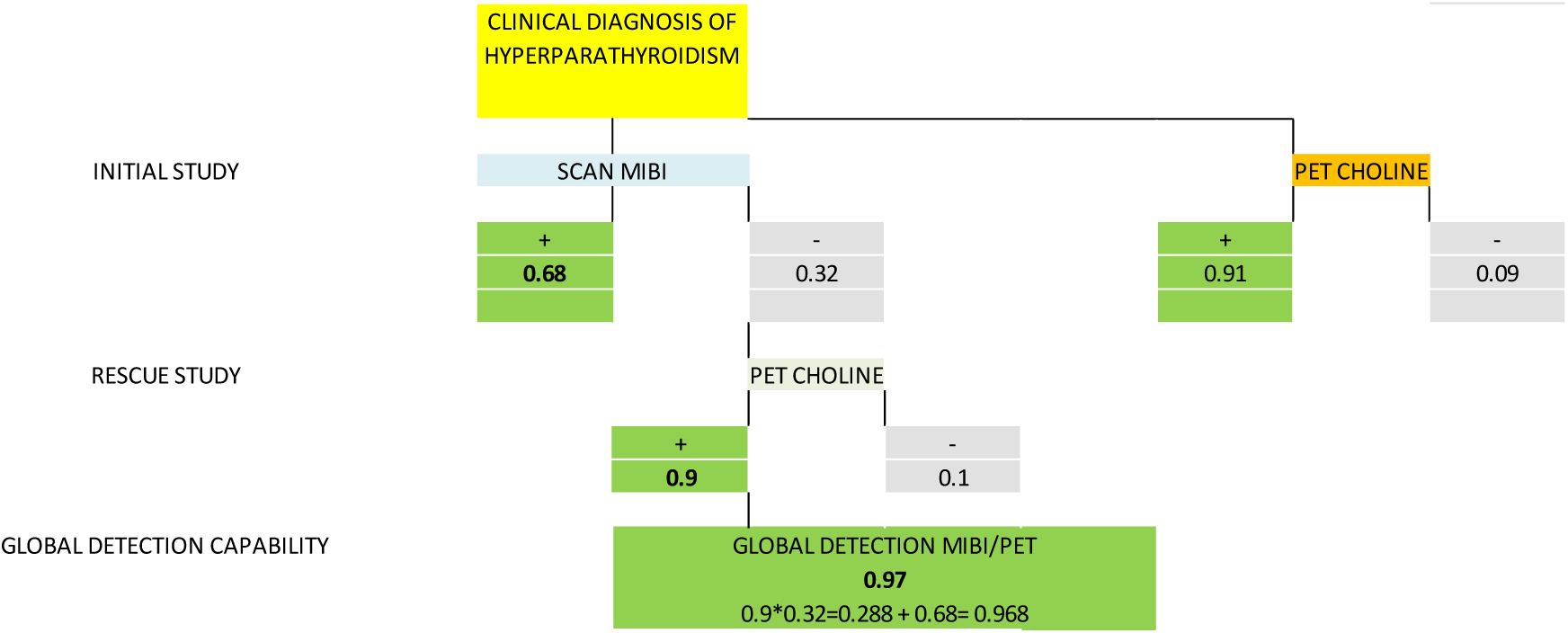
Prevalence of uptake with SPECT/TC-MIBI and PET/TC-Choline in hyperparathyroidism.

The CH localization study using PET/TC offers advantages such as higher number of uptakes (both the number of patients with at least one uptake and the higher number of multiple adenoma or hyperplasia captures [20], shorter imaging time [14] and exposure of patients with less radiation than the SPECT/CT hybrid image with Tc-99m-sestaMIBI [50].

As disadvantages is the difficulty in distinguishing catches in thyroid, nodes, thymus or muscle (false positives) [50] and logistical problems [14].

CH scan may be indicated both as an initial scan and in patients with prior negative MIBI study.

## Conclusions

1. Ultrasound-associated SPECT/TC-MIBI scan is currently the scan of choice for the location of pathological parathyroid glands prior to surgery.
2. PET-Choline scan increases the number of patients with at least one uptake on both the first scan and on scans with initial negative MIBI studies.
3. Currently PET-Choline scan seems an option to locate pathological glands in patients with negative initial scan.

## Data Availability

All data are published in the article

